# Value of the body roundness index vs. body mass index for predicting 9-year mortality: *Shizuoka Kokuho Database* study

**DOI:** 10.64898/2025.12.19.25342719

**Authors:** Masato Takeuchi

## Abstract

**Background:** The body roundness index (BRI), calculated from height and waist circumference, is a novel obesity metric proposed as an alternative to body mass index (BMI). Although the BRI is reportedly associated with mortality risk, little is known about how the performance of the BRI and BMI compare.

**Methods:** We used data from the SKDB, a regional healthcare database in Shizuoka, Japan, that includes data from >2 million people from 2012 to 2022. Individuals aged 40 to 74 years who received a Specific Health Checkups in 2013 were included and followed for up to nine years. We investigated the associations of the BRI and BMI with future risk of all-cause death and compared their predictive value using the net reclassification index (NRI) and c-statistics.

**Results:** Among 202,133 individuals (median age: 66 years; 64.6% female; median BMI: 22.3 kg/m²), 10,336 deaths (5.2%) occurred over a median follow-up of eight years. Both the BRI and BMI demonstrated J-shaped associations with mortality risk. Incorporating the BRI or BMI into age- and sex-adjusted models improved NRI and c-statistics, with NRI increases of 5.6% (95% confidence interval [CI]: 3.6 to 7.1%) for BRI and 7.8% (95% CI: 6.0 to 9.6%) for BMI. The incremental improvements in the c-statistic were 0.48% (95% CI, 0.47 to 0.48%) for the BRI and 0.65% (95% CI, 0.64 to 0.65%) for BMI, whereas the BRI performed slightly worse than BMI (difference: –0.17%; 95% CI, –0.16 to –0.18%).

**Conclusions:** Although BRI was predictive of mortality risk, the BRI did not outperform BMI in our population.

## Introduction

Overweight and obesity (hereafter referred to as overweight/obesity for simplicity) are endemic, with increasing rates observed both globally and regionally^1^. With this continuing trend, more than half of the adult population worldwide is projected to be overweight/obese by 2050^2^.

Overweight/obesity is often measured by body mass index (BMI) or its categorization: normal weight for a BMI of ≥ 18.5 kg/m^2^ to < 25.0 kg/m^2^, overweight for a BMI of ≥ 25.0 kg/m^2^ to < 30 kg/m^2^, and obesity for a BMI of ≥ 30.0 kg/m^2^ ^3^. Although conventional and well recognized, BMI has been criticized as it does not adequately represent body composition, such as adiposity^4,5^. In response to this inherent limitation, alternative metrics have been proposed to better characterize anthropometric indices of adiposity^6^. The body roundness index (BRI), proposed in 2013, is among these metrics^7^. The BRI is calculated from height waist circumference (WC) based on the premise that the human body’s shape, particularly in the context of obesity, can be approximated as an ellipse. It was recently reported that the BRI is associated with mortality and obesity-related non-communicable disease^8–11^. However, data concerning the comparative utility between the BRI and BMI are limited, particularly in longitudinal, population-based settings.

We therefore compared the BRI and BMI in terms of their ability to predict mortality by leveraging a regional healthcare database in Japan.

## Materials and methods

### Data source and study population

We used municipal-level health insurance data in Shizuoka Prefecture, Japan (Shizuoka Kokuho Database [SKDB]), covering data from self-employed and unemployed individuals (including retirees). The cohort profile of the SKDB has been published elsewhere^12^. The database has been updated regularly since that publication^13^, and the version used in this study (SKDB 2024) included approximately two million persons from April 2012 to September 2022. All data in the SKDB were anonymized.

The SKDB contains records from multiple sources, including insurance claims, annual health checkup data (specifically, data from Specific Health Checkups [SHC], which are offered primarily to all residents aged 40 to 74 years), residential information, and death records, where applicable. All records were linked by a unique identifier throughout.

This study utilized data from SHC recipients in 2013. Information on height, body weight, and WC from SHCs were routinely collected, as were information on self-reported smoking status, alcohol intake, and regular exercise^14^. WC was measured at the umbilical level according to standardized SHC procedures^15^, and all measurements were performed by trained staff (e.g., nurses). Information on comorbid conditions was obtained from medical insurance claims before SHC receipt. To ensure data accuracy, participants were required to have at least 12 months of continuous inclusion in the SKDB prior to undergoing a SHC for study inclusion.

Data cleaning, such as the treatment for outliers, was performed before formal analysis (Supplemental text).

This study was conducted under the approval of our Institutional Review Borad (SGUPH_2021_001_096, approved on September 3rd, 2024). As all data were anonymized, informed consent from each enrollee was waived.

### Exposure and outcome

The primary exposure of interest was the BRI in 2013, estimated using data in the SHC setting. The BRI was calculated with the following equation^7^:

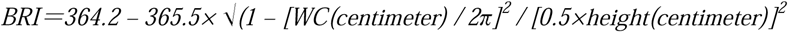

BMI was calculated as body weight (in kilograms) divided by height squared (in meters) using the checkup data.

The outcome was all-cause death, recorded in the SKDB along with the date of death.

### Statistical analysis

We summarized participant characteristics using descriptive statistics and reported them where appropriate.

We first examined whether the BRI could predict future mortality risk. The probability of all-cause mortality at nine years was estimated using a Cox regression model, where the BRI was categorized into 0.1-unit bins in an age- and sex-standardized population^16^. In this model, we expected a nonlinear relationship between the BRI and mortality^8^; therefore, the BRI was converted into a restricted cubic spline with four knots^17^. Follow-up began from the receipt of the checkup day in 2013 and continued until the earliest event of death, disenrollment from health insurance, or the administrative database end of September 2022. The same analysis was conducted for BMI, categorized into 0.5-unit bins. We then created plots displaying the relationships between each index (BRI or BMI) and the predicted probability of all-cause mortality.

Furthermore, we developed three models to predict mortality: a base model adjusted for age and sex; a BRI model adjusted for age, sex, and BRI (spline function); and a BMI model adjusted for age, sex, and BMI (spline function). Predictive performance was calculated using the net reclassification index (NRI) to assess risk stratification^18^ and c-statistics to measure model discrimination between the BRI and BMI^19^. A 95% confidence interval (CI) for the NRI and c statistic was estimated using 1,000 bootstrap replicates.

All statistical analyses were performed in R version 4.5.1 (https://cran.r-project.org/) with the help of its distributed packages (Supplemental text).

### Sensitivity analyses

Three sensitivity analyses were conducted. First, full models were constructed that further adjusted for smoking status, alcohol intake, regular exercise, municipal-level area deprivation index^20^, and Charlson comorbidity index^21^. Missing data on smoking status, alcohol intake, and regular exercise (Table 1) were addressed using multiple imputation with 50 imputations and the chained equations approach^22^; we assumed that the missingness pattern was random. Second, age- and sex-standardization was performed to match the Japanese population aged 40 to 74 years. The population structure was obtained from the 2015 national census^23^, in which age was categorized into 5-year bins. Third, the BRI and BMI were categorized into binary groups, and hazard ratios (HRs) were calculated. Although there is a standard cutoff for BMI (e.g., normal weight defined as a BMI of 18.5–<25.0), there is no consensus regarding the optimal cutoff point for the BRI. Therefore, we adopted a data-driven approach, and the cutoff point was determined using the Youden index^24^. BMI was classified into two categories—18.5–<25.0 kg/m^2^ (normal weight, reference) and ≥25.0 kg/m^2^ (overweight/obesity)—to ensure consistency with the binary categorization of BRI. To align with this categorization, individuals with a BMI <18.5 were excluded from this sensitivity analysis.

**Table 1:** Characteristics of the final study cohort (N=200,133)

| **Variable Name** | **Value<sup>1</sup>** | **Missing (if any)** |
| --- | --- | --- |
| ***Age (yrs)*** | 66 (62-70) | NA |
| ***Sex*** | Female: 129,386 (64.6%) | NA |
| ***BMI (kg/m<sup>2</sup>)*** | 22.3 (20.3, 24.5) | NA |
| ***categorical*** | Underweight: 17,613 (8.8%)<br>Normal weight: 140,615 (70%)<br>Overweight: 36,591 (18%)<br>Obesity: 5,314 (2.7%) |  |
| ***BRI*** | 3.69 (2.98, 4.52) | NA |
| ***waist circumstance (cm)*** | Male: 84.0 (79.0, 89.2)<br>Female: 80.0 (74.0, 86.6) | NA |
| ***Smoking*** | Yes: 22,519 (11%) | 45 (0.02%) |
| ***Alcohol intake*** | Regularly: 34,800 (20%)<br>Occasionally: 36,416 (21%)<br>Raley/never: 106,034 (60%) | 22, 883 (11.4%) |
| ***Regular exercise*** | Yes: 74,350 (43%)<br>No: 98,770 (57%) | 27.013 (13.5%) |

1: number (with frequency) or median (with interquartile range)

## Results

### Characteristics of the study cohort

The study cohort included 200,133 individuals (Table 1). The median age (interquartile range [IQR]) was 66 (62–70) years, and 64.6% of the participants were female. The median BMI and BRI were 22.3 (IQR, 20.3–24.5) and 3.69 (IQR, 2.98–4.52), respectively.

Over a median follow-up of eight years, 10,336 individuals (5.2%) had died.

### Primary analysis

The relationships of the BRI and BMI with all-cause mortality are presented in Figures 1 and Figure 2. Both indices demonstrated a J-shaped relationship, with the lowest mortality risk observed at a BRI of 3.7 and a BMI of 22.5 kg/m².

**Figure 1.**
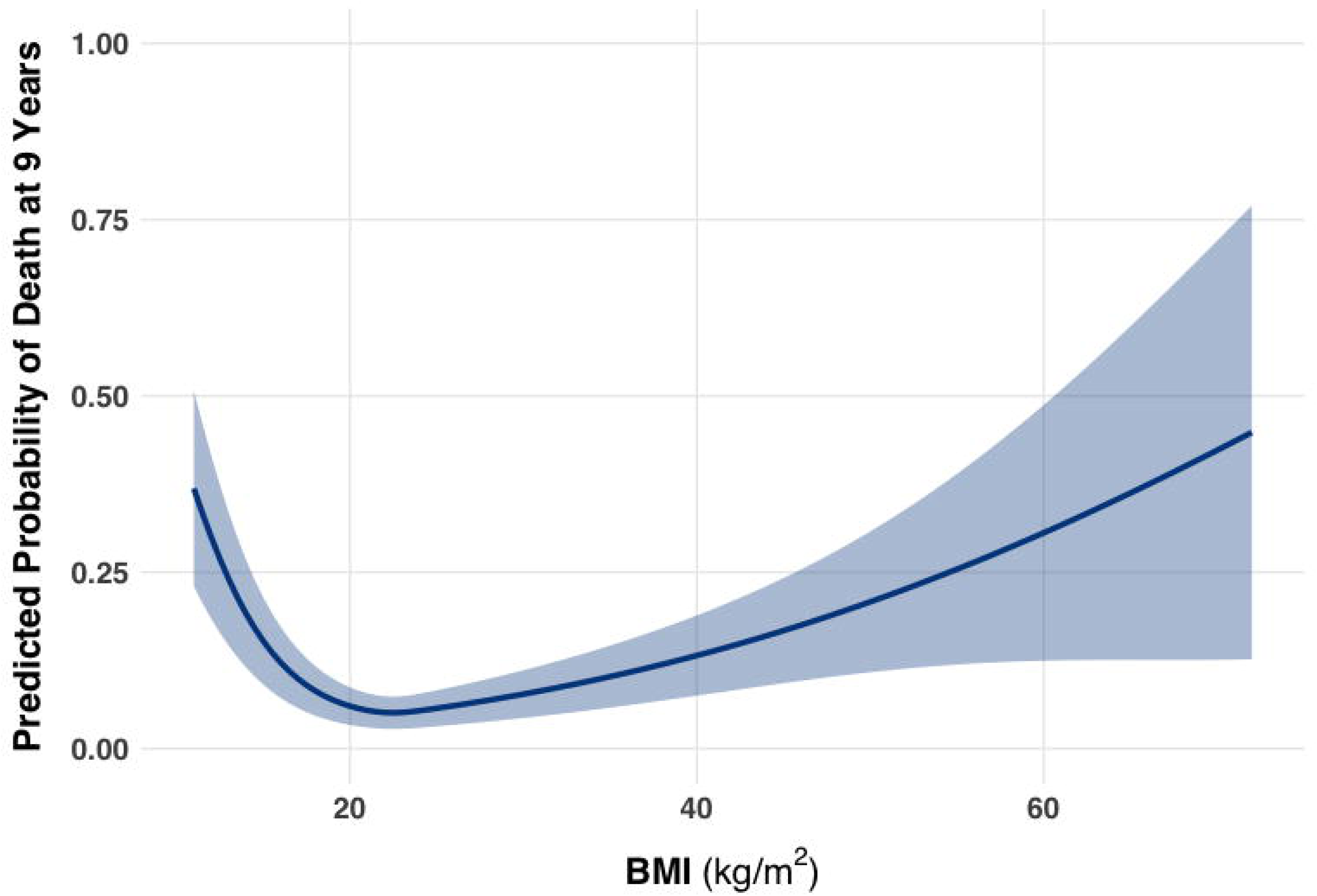

**Figure 2.**
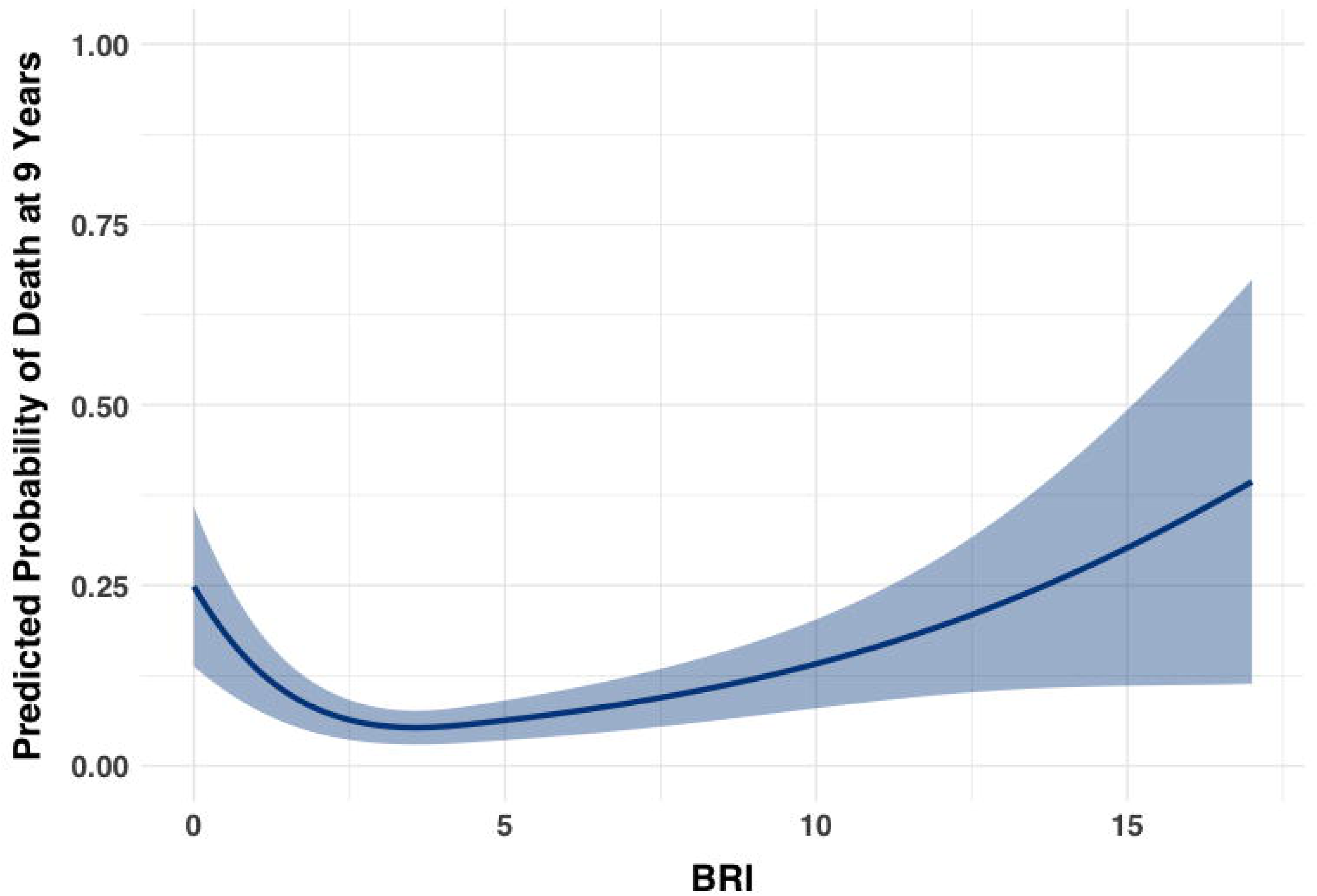

Compared with the age- and sex-adjusted model (base model), both the NRI and c-statistics improved in models further adjusted for the BRI (BRI model) or BMI (BMI model). The NRIs for the base model vs. the BRI model and the base model vs. the BMI model were 5.7% (95% CI, 3.8–7.3%) and 7.9% (95% CI, 5.9–9.7%), respectively, with overlapping 95% CIs. The c-statistics for the base, BRI, and BMI models were 70.3% (95% CI, 70.3–70.3%), 70.8% (95% CI, 70.8–70.8%), and 71.0% (95% CI, 70.9–71.0%), respectively. Differences in c-statistics were 0.48% (95% CI, 0.48–0.49%) for the base model vs. the BRI model and 0.66% (0.65–0.66%) for the base model vs. the BMI model. In contrast, the difference in the c-statistic between the BMI and BRI models was –0.17% (95% CI, –0.16 to –0.18%), indicating the poorer discrimination performance of the BRI relative to the BMI.

### Sensitivity analysis

In the first sensitivity analyses, which were fully adjusted for lifestyle factors and comorbidities, the results were essentially identical to those in the primary analysis. The relationships of the BRI and BMI with all-cause mortality showed a J-shaped pattern, with the lowest mortality risk observed at a BRI of 3.7 and a BMI of 22.5 kg/m² (Supplemental Figures 1 and 2). The confidence bands were somewhat wider, possibly due to increased uncertainty arising from missing data for some variables. The NRI and c-statistic results are summarized in Supplemental Table 1; the findings were similar to those in the primary analysis except for an approximately 2% improvement in the c-statistic.

Second, the population age and sex structure was standardized across all of Japan, with a median age of 55–59 years, and females accounted for 49.9%. As in the primary analysis, the risk was J-shaped (Supplemental Figures 3 and 4), with the lowest risk at 3.5 for BRI and 22.5 kg/m² for BMI. Although the NRI and c-statistic differed slightly from those in the primary analysis (Supplemental Table 2), the overall results were unchanged; the BRI was not superior to BMI for mortality risk prediction.

Finally, the BRI was dichotomized at a cutoff value of 4.9, as determined by the Youden index. In this analysis involving 184,857 individuals with a BMI ≥ 18.5 kg/m², individuals in the higher BRI group had an increased risk of mortality, with an HR of 1.38 (95% CI: 1.33–1.43). This association was more pronounced than the estimated risk from dichotomized BMI, with an HR of 1.17 (95% CI: 1.12–1.22), a cutoff of 25.0 kg/m², or an HR of 1.11 (95% CI: 1.07–1.15), and a Youden index-based cutoff of 24.1 kg/m².

## Discussion

In our study, both the BRI and BMI were able to predict mortality risk. However, the predictive performance of the BRI was no better than that of the BMI, except in one sensitivity analysis in which the BRI was exploratory grouped into two categories using a cutoff of 4.9. As a novel anthropometric index, the BRI has attracted increasing attention (Supplemental Figure 5). Notably, it has been reported to be associated with mortality^8^. However, few studies have evaluated the performance of the BRI relative to BMI in predicting mortality risk using longitudinal cohort data. As of September 2025, a PubMed search using the keyword “body roundness index” retrieved nearly 550 citations of all types, including review articles, commentaries, and preprints.

Among these, 24 original studies examined the association between the BRI and subsequent all-cause or cause-specific mortality. Of these, only two articles directly compared the performance of the BRI and BMI for mortality risk—one for cardiovascular mortality^25^ and another for all-cause mortality^26^. Among these two studies, Ichikawa et al. evaluated the performance of the BRI relative to BMI, with cardiovascular death as one component of a composite outcome; however, results solely for mortality were not reported. In another study using UK Biobank data, Mäkinen and colleagues compared the BRI with BMI alone (or with the combination of BMI and waist-to-hip ratio) and reported that the BRI was a better predictor of mortality. However, as the authors noted, the generalizability of their findings may be limited because most participants were of European descent, and body shape differs across populations. Therefore, evidence is currently limited on the relative performance of the BRI with respect to mortality risk. Our study addresses this gap by evaluating the comparative performance of the BRI and BMI in predicting mortality risk in a Western Asian population.

BMI is often categorized into normal weight (18.5–<25 kg/m²), overweight (25–<30 kg/m²), and obesity (≥30 kg/m²). Such categorization—along with intuitive labels—may be beneficial, as it can improve interpretability and facilitate communication of obesity status^27^. In our study, when the BRI was binary categorized using the Youden index, obesity (defined as a BRI >4.6) was associated with mortality (HR: 1.38), whereas the HR was 1.17 or 1.11 when categorized BMI was used. These findings suggest that obesity defined by the categorized BRI was associated with a more pronounced risk of death than obesity classified by BMI. However, these results should be interpreted with caution for the following reasons. First, the cutoff point was determined in a data-driven manner and may be population-specific, thereby limiting generalizability. Second, the optimal cutoff may vary depending on the outcome of interest, such as the development of noncommunicable diseases (e.g., type 2 diabetes). Finally, it remains unclear whether categorizing the BRI in this manner actually improves its clinical utility, including increased recognition of obesity^28^. Therefore, further validation is needed to determine whether categorical BRI offers any advantage over standard BMI categorization.

There are several limitations to our study. First, the generalizability of our findings may be limited, as our study population was derived from enrollees in a regionally based insurance plan who participated in the SHC in 2013. To partially address this limitation, we conducted a sensitivity analysis in which the study sample was reweighted to match the age and sex distribution of the entire Japanese population. In this analysis, the reweighted population was approximately 10 years younger and had a more balanced sex ratio than the original study population; however, the primary results remained unchanged. Nevertheless, overweight/obesity was less prevalent in our study cohort; the median BMI was 22.3 kg/m^2^, and individuals with overweight/obesity accounted for 20.9% of the participants. Therefore, the results may differ in populations with a higher prevalence of obesity. Second, although WC was measured at the umbilical level during a standardized SHC, there are some variations in the measurement site, such as the midpoint between the lowest rib and the iliac crest^29^. However, one systematic review found that WC measurement protocols do not substantially influence the association between WC and all-cause mortality^30^. Therefore, we assumed that differences in the WC measurement methods were unlikely to materially affect our findings. Third, we were unable to identify the cause of death in our database. Thus, a subset of observed deaths may be unrelated to obesity.

## Conclusions

Our findings suggest that in the Japanese population, the BRI does not offer clear advantages over BMI for predicting all-cause mortality. Further studies in diverse populations and across different obesity-related outcomes are warranted to clarify whether the BRI provides incremental clinical utility beyond established anthropometric measures—BMI.

## Data Availability

Data distribution is not permitted due to the contract with data provider.

## References

1. NCD Risk Factor Collaboration (NCD-RisC). Worldwide trends in underweight and obesity from 1990 to 2022: a pooled analysis of 3663 population-representative studies with 222 million children, adolescents, and adults. Lancet. 2024;403:1027–1050.

2. GBD 2021 Adult BMI Collaborators. Global, regional, and national prevalence of adult overweight and obesity, 1990-2021, with forecasts to 2050: a forecasting study for the Global Burden of Disease Study 2021. Lancet. 2025;405:813–838.

3. González-Muniesa P, Mártinez-González MA, Hu FB, et. al. Obesity. Nat Rev Dis Primers. 2017;3:17034.

4. Bray GA. Beyond BMI. Nutrients. 2023;15:2254.

5. Rubino F, Cummings DE, Eckel RH, et. al. Definition and diagnostic criteria of clinical obesity. Lancet Diabetes Endocrinol. 2025;13:221–262.

6. Piqueras P, Ballester A, Durá-Gil JV, Martinez-Hervas S, Redón J, Real JT. Anthropometric Indicators as a Tool for Diagnosis of Obesity and Other Health Risk Factors: A Literature Review. Front Psychol. 2021;12:631179.

7. Thomas DM, Bredlau C, Bosy-Westphal A, et. al. Relationships between body roundness with body fat and visceral adipose tissue emerging from a new geometrical model. Obesity (Silver Spring). 2013;21:2264–2271.

8. Zhang X, Ma N, Lin Q,et. al. Body Roundness Index and All-Cause Mortality Among US Adults. JAMA Netw Open. 2024;7:e2415051.

9. Kawasoe S, Kubozono T, Salim AA, et. al. Association between anthropometric indices and 5-year hypertension incidence in the general Japanese population. Hypertens Res. 2024;47:867–876.

10. Qiu L, Xiao Z, Fan B, Li L, Sun G. Association of body roundness index with diabetes and prediabetes in US adults from NHANES 2007-2018: a cross-sectional study. Lipids Health Dis. 2024;23:252.

11. Khanmohammadi S, Fallahtafti P, Habibzadeh A, et. al. Effectiveness of body roundness index for the prediction of nonalcoholic fatty liver disease: a systematic review and meta-analysis. Lipids Health Dis. 2025;24:117.

12. Nakatani E, Tabara Y, Sato Y, Tsuchiya A, Miyachi Y. Data Resource Profile of Shizuoka Kokuho Database (SKDB) Using Integrated Health- and Care-insurance Claims and Health Checkups: The Shizuoka Study. J Epidemiol. 2022;32:391–400.

13. KDB Committee, Shizuoka Graduate University of Public Health. Shizuoka Kokuho Database (SKDB). https://skdb.s-sph.ac.jp/en; Accessed 14.12.2025.

14. Iseki K, Asahi K, Moriyama T, Yamagata K, Tsuruya K, Yoshida H, Fujimoto S, Konta T, Kurahashi I, Ohashi Y, Watanabe T. Risk factor profiles based on estimated glomerular filtration rate and dipstick proteinuria among participants of the Specific Health Check and Guidance System in Japan 2008. Clin Exp Nephrol. 2012;16:244–249.

15. Hu H, Kawasaki Y, Kuwahara K, et. al. Trajectories of body mass index and waist circumference before the onset of diabetes among people with prediabetes. Clin Nutr. 2020;39:2881–2888.

16. Sjölander A. Regression standardization with the R package stdReg. Eur J Epidemiol. 2016;31:563–574.

17. Discacciati A, Palazzolo MG, Park JG, Melloni GEM, Murphy SA, Bellavia A. Estimating and presenting non-linear associations with restricted cubic splines. Int J Epidemiol. 2025;54:dyaf088.

18. Leening MJ, Vedder MM, Witteman JC, Pencina MJ, Steyerberg EW. Net reclassification improvement: computation, interpretation, and controversies: a literature review and clinician’s guide. Ann Intern Med. 2014;160:122–131.

19. Pencina MJ, D’Agostino RB Sr. Evaluating Discrimination of Risk Prediction Models: The C Statistic. JAMA. 2015;314:1063–1064.

20. Nakaya T, Honjo K, Hanibuchi T, et.al. Associations of all-cause mortality with census-based neighbourhood deprivation and population density in Japan: a multilevel survival analysis. PLoS One. 2014;9:e97802.

21. Quan H, Li B, Couris CM, et. al. Updating and validating the Charlson comorbidity index and score for risk adjustment in hospital discharge abstracts using data from 6 countries. Am J Epidemiol. 2011;173:676–682.

22. Blazek K, van Zwieten A, Saglimbene V, Teixeira-Pinto A. A practical guide to multiple imputation of missing data in nephrology. Kidney Int. 2021;99:68–74.

23. Statistics Bureau of Japan. 2015 Population Census. https://www.e-stat.go.jp/en/stat-search/files?page=1&layout=datalist&toukei=00200521&tstat=000001080615&cycle=0&tclass1=000001124175&cycle_facet=tclass1&tclass2val=0; Accessed 14.12.2025.

24. Raghavan R, Ashour FS, Bailey R. A Review of Cutoffs for Nutritional Biomarkers. Adv Nutr. 2016;7:112–120.

25. Ichikawa T, Okada H, Nakajima H, et.al. Anthropometric indices for predicting incident cardiovascular disease: a 13-year follow-up study in a Japanese population. Am J Clin Nutr. 2025;122:954–961.

26. Mäkinen VP, Zhao S, Ihanus A, Tynkkynen T, Ala-Korpela M. Epidemiological associations between obesity, metabolism and disease risk: are body mass index and waist-hip ratio all you need? Int J Obes (Lond). 2025;49:2555–2560.

27. Royston P, Altman DG, Sauerbrei W. Dichotomizing continuous predictors in multiple regression: a bad idea. Stat Med. 2006;25:127–141.

28. Gee KA, Thompson HR, Sliwa SA, Madsen KA. BMI Reporting and Accuracy of Child’s Weight Perception. Pediatrics. 2022;150:e2021055730.

29. Ness-Abramof R, Apovian CM. Waist circumference measurement in clinical practice. Nutr Clin Pract. 2008;23:397–404.

30. Ross R, Berentzen T, Bradshaw AJ, et. al. Does the relationship between waist circumference, morbidity and mortality depend on measurement protocol for waist circumference? Obes Rev. 2008;9:312–325.

