## Supplemental files for "Value of the body roundness index vs. body mass index for predicting 9-year mortality: *Shizuoka Kokuho Database* study"

- Statistical environment
- Data cleaning
- Sensitivity analyses

**Statistical environment**

- stdReg2 (version, 1.0.3): Risk standardization
- riskRegression (version, 2025.09.17): time‐dependent receiver operating characteristic calculation
- survIDINRI (version, 1.1-2): time‐dependent net reclassification improvement calculation
- mice (version, 3.18.0): multiple imputation in the sensitivity analysis

(Other commonly-used packages such as base or survival are not listed here.)

**Data cleaning**

We referred to the research on worldwide trends in underweight and obesity^1^, and pre-specified the outliner for body mass index (BMI, kg/m^2^), height (cm), weight (kg), and waist circumstance (WC, cm):

- BMI ≥ 80 OR BMI ≤10
- height ≥ 250 OR height ≤ 100
- weight ≥ 300 OR weight ≤ 12
- WC ≥ 200 OR WC ≤ 30

Individuals who had at least one record of outliner were excluded from our study.

In contrast to BMI, the outlier definition for the body roundness index (BRI) is not well established. In the original study by Thomas^2^, BRI values ranged from 1 to 16 in the US population; however, this range may vary across populations. Therefore, in our study, we did not specify any outliers for BRI if BRI value was calculable and greater than 0.

**Reference**

1. NCD Risk Factor Collaboration (NCD-RisC). Worldwide trends in underweight and obesity from 1990 to 2022: a pooled analysis of 3663 population-representative studies with 222 million children, adolescents, and adults. *Lancet.* 2024 Mar 16;403(10431):1027-1050. doi: 10.1016/S0140-6736(23)02750-2.
2. Thomas DM, Bredlau C, Bosy-Westphal A, Mueller M, Shen W, Gallagher D, Maeda Y, McDougall A, Peterson CM, Ravussin E, Heymsfield SB. Relationships between body roundness with body fat and visceral adipose tissue emerging from a new geometrical model. *Obesity (Silver Spring)*. 2013 Nov;21(11):2264-71. doi: 10.1002/oby.20408.

**Sensitivity Analysis 1: Multiple Imputation–Based Full Covariate Model**

**Supplemental Figure 1.** Association of BMI with the risk of all-cause mortality at 9 years

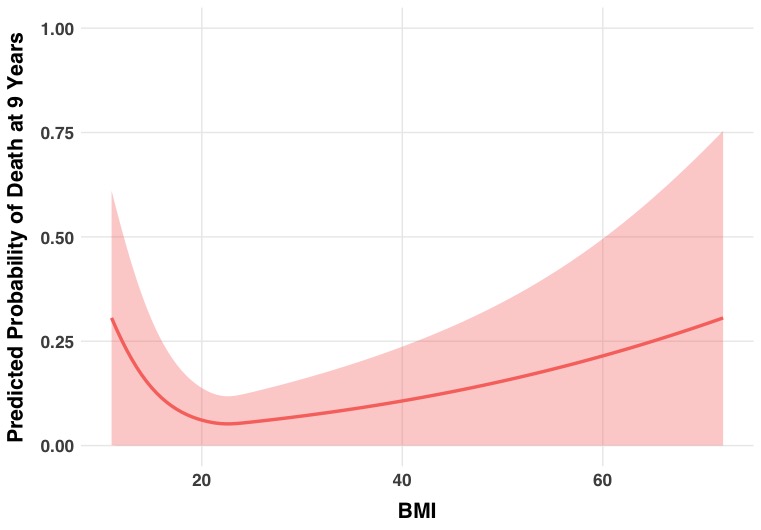

BMI: body mass index (kg/m^2^)

**Supplemental Figure 2.** Association of BRI with the risk of all-cause mortality at 9 years

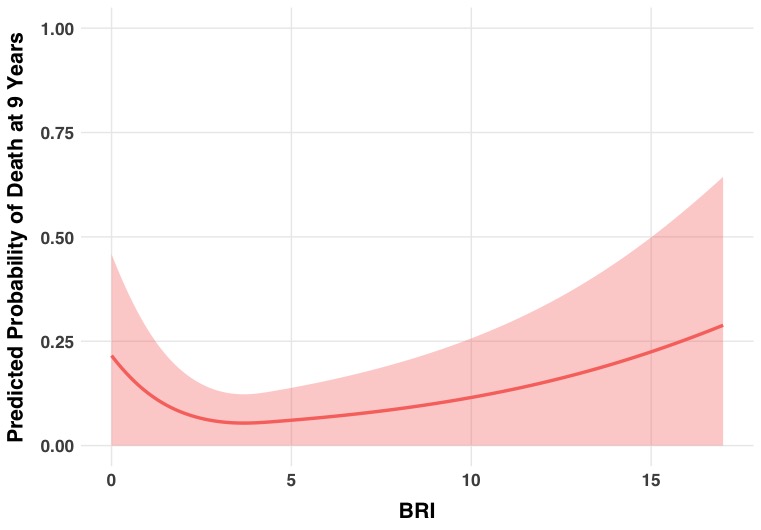

BRI: body roundness index

**Supplemental Table 1**. Comparison of Net Reclassification Index

| **Comparison** | **Difference (95% confidence interval)** |
| --- | --- |
| **Model with vs without BMI** | 6.9% (4.8%, 8.5%) improvement |
| **Model with vs without BRI** | 4.7% (2.9%, 6.5%) improvement |
| **Model with BMI vs with BRI** | -4.1% (-6.3%, -1.8%) decrease (in the BRI model) |

**Supplemental Table 2**. Comparison of c-statistics

| **Comparison** | **Difference (95% confidence interval)** |
| --- | --- |
| **Model with vs without BMI** | 72.5% vs 72.9%  ^­^0.40 % (0.40%, 0.40%) improvement |
| **Model with vs without BRI** | 72.5% vs 72.8%  0.28% (0.28%, 0.28%) improvement |
| **Model with BMI vs with BRI** | 72.9% vs 72.5%  -0.12% (-0.12%, -0.12%) decrease |

**Sensitivity Analysis 2: Age- and Sex-Standardized to the Population Structure of Japan**

**Supplemental Figure 3.** Association of BMI with the risk of all-cause mortality at 9 years

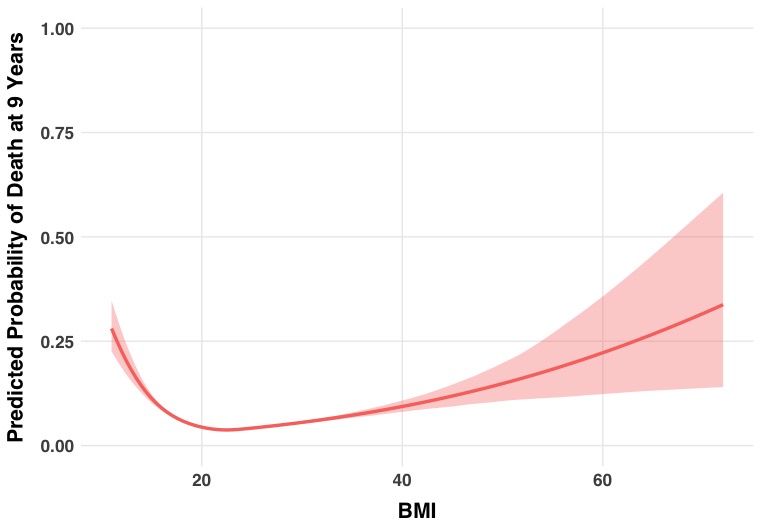

BMI: body mass index (kg/m^2^)

**Supplemental Figure 4**: Association of BRI with the risk of all-cause mortality at 9 years

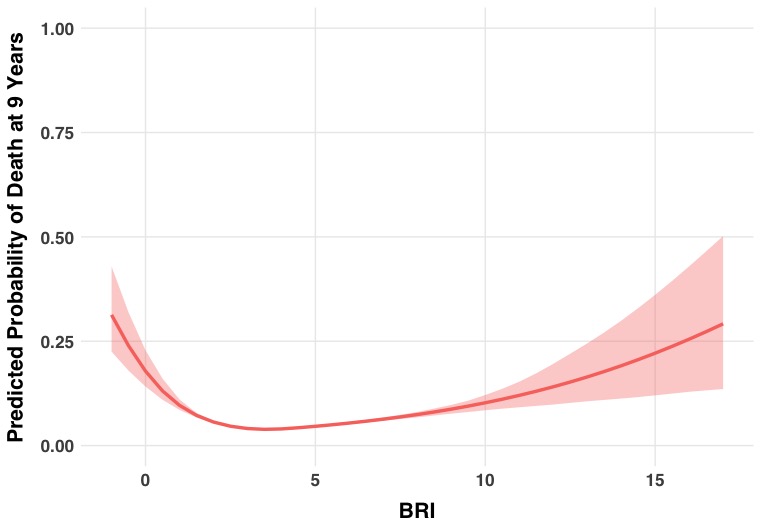

BRI: body roundness index

**Supplemental Table 3**. Comparison of Net Reclassification Index

| **Comparison** | **Difference (95% confidence interval)** |
| --- | --- |
| **Model with vs without BMI** | 7.9% (6.0%, 9.6%) improvement |
| **Model with vs without BRI** | 6.0% (4.4%, 7.5%) improvement |
| **Model with BMI vs with BRI** | -0.11% (-6.1%, 4.7%) decrease (in the BRI model) |

**Supplemental Table 4**. Comparison of c-statistics

| **Comparison** | **Difference (95% confidence interval)** |
| --- | --- |
| **Model with vs without BMI** | 69.8 % vs 70.6%  ^­^0.88 % (0.87%, 0.89%) improvement |
| **Model with vs without BRI** | 69.8% vs 70.5%  0.75% (0.74%, 0.75%) improvement |
| **Model with BMI vs with BRI** | 70.6% vs 70.5%  -0.13% (-0.14%, -0.12%) decrease |

**Supplemental Figure 5: Number of publications on body roundness index**

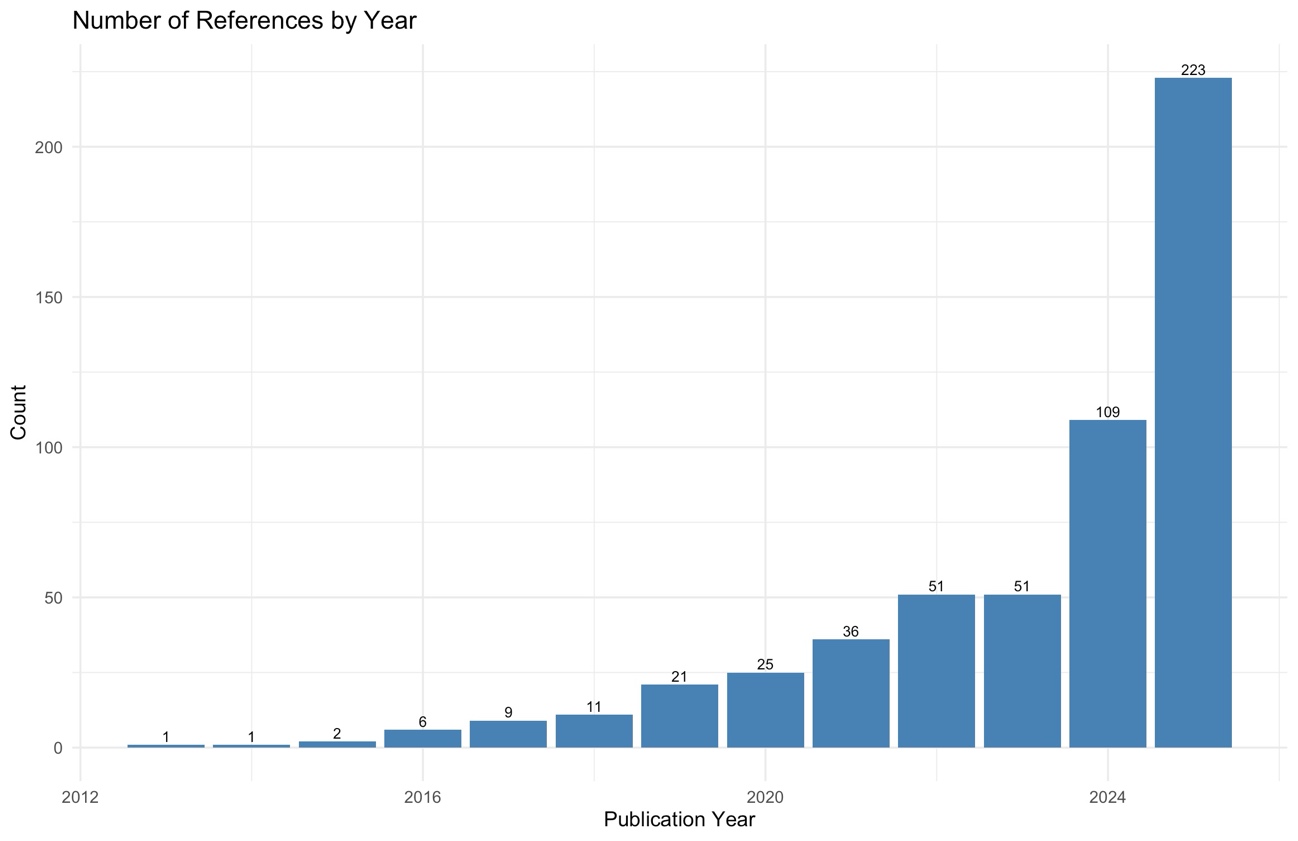
